# Co-designing age-specific meal plans using locally available foods for under-five children in Tanzania: A human-centered design protocol

**DOI:** 10.1101/2025.02.19.25322530

**Authors:** Aminieli Itaeli Usiri, Samson Charles, Aneth Gabriel Mhidze, Jackline Estomihi Kiwelu

## Abstract

**Background:** Infant and young child nutrition is fundamental to individual health and national economic growth. Malnutrition remains a major challenge, particularly in developing countries such as Tanzania. Despite being food-producing regions, Njombe and Iringa continue to experience high rates of undernutrition among children under five, largely due to knowledge gaps and poor feeding practices among caregivers.

**Aim:** To co-design age-specific meal plans using locally available foods in Njombe and Iringa, Tanzania. Research seeks to develop culturally, and economically feasible meal plans to improve child nutrition in Tanzania.

**Methods:** A five-step Human centered design approach will be employed: (1) **Exploration** through focus group discussions and key informant interviews to understand caregivers feeding challenges; (2) **Co-design** to develop meal plans incorporating affordable, locally available; (3) **Validation and Refinement** through focus group discussion and stakeholder feedback; (4) **Implementation** with trained Community Health Workers supporting caregivers in the adoption of meal plan; and (5) **Evaluation** using qualitative and quantitative methods to assess effectiveness and adoption over a one-year period. A total of 208 participants will be purposefully selected. Thematic analysis will be conducted using NVivo 12 software to identify key challenges caregivers face, facilitators, and locally available and affordable foods to be used in co-design of meal plan.

**Expected results:** The findings will provide insights about feeding challenges caregivers of under five children faces, meal plan prototypes tested and scaled up if proven successful. Successful implementation of these age specific meal plans, this study anticipates improved caregiver knowledge and practices regarding infant and young child feeding, increased dietary diversity, and enhanced nutritional outcomes among children under five children in Tanzania.

## Introduction

Infant and young child nutrition is vital for individual health and a nation’s economic growth (1). Childhood nutrition serves as the cornerstone of a lively and thriving nation. According to United Nations International Children’s Emergency Fund (UNICEF) good nutrition is the bedrock of the children survival and development (2). Undernutrition hinders children ability to learn, grow healthy, earn as adults, and participate meaningfully in their communities (2, 3). Moreover, their immunity system is weakened, and face increased risk of diseases and death (4). Globally in 2022, 149 million children under 5 were estimated to be stunted (too short for age), while 45 million were estimated to be wasted (too thin for height) (5). These cases have contributed to nearly half deaths of under-five children worldwide, while most of them coming from lower and middle income countries (5). Tanzania being among lower- and middle-income countries still suffers higher prevalence of undernutrition and its associated effects. The demographic health survey of 2022 indicated that 30% of under-five children are stunted, 9% severely stunted, 3% are wasted, and 12% are underweight. The survey further indicates that only 1 child in 23 children survive to their fifth birthday (6). Unfortunately these high prevalence are from regions that produce more food in the country namely: Njombe, Iringa, Rukwa and Ruvuma (7, 8).

UNICEF highlighted factors that causes childhood undernutrition including low food intake, infection, including financial constraints, limited parental education, and inadequate feeding practices (9). Proper infant and young child feeding (IYCF) practices, tailored to a child’s age, is essential for ensuring adequate nutrition (10). Studies show that for children aged 6 to 23 months, the meal plan should incorporate a diverse array of foods that not only meet their energy requirements but also provide essential nutrients for growth and cognitive development (10, 11). The emphasis on dietary diversity is crucial, as it has been shown to enhance overall nutrient intake and improve health outcomes (12). Furthermore, regular feeding schedules that align with the recommended meal frequencies will help establish healthy eating habits in early age (13). However, in Tanzania, knowledge on feeding practices among women or caregivers is far beyond the recommended levels by Food and Agricultural Organization (14). Studies done in rural Tanzania reported a significant portion of the population had inadequate knowledge and practices related to nutrition. For instance, only 14% of mothers or caregivers reported receiving nutrition education and optimal practices. Consumptions of fruits and poultry were minimally practiced at 22% and 2% respectively (14, 15). Walter C, et al (16) who assessed feeding practices, dietary diversity and dietary adequacy among caregivers of under-five children in Dodoma, showed that 66.1% of caregivers had unsatisfactory feeding practices, 67.8% feed their children inadequately and only 32.2% of caregivers attained minimum dietary diversity to their children. Infants and young children suffer the consequences of this knowledge and practices barriers and end up in undernutrition as reported by Masuke et al (17).

Attaining Sustainable Development Goal 3, which focuses on ensuring healthy lives and promoting well-being for all ages, demands increased efforts to overcome knowledge barriers and improve feeding practices. Several interventions such as Addressing Stunting in Tanzania (ASTUTE) project, school feeding programmers’, *Stawisha Maisha* - *Nourishing Life* program in Tanzania and many others have been implemented and improvements have been observed (18–20). Despite the observed improvements the country failed to achieve its national nutritional plan of reducing stunted population to 28% by 2021(18). A notable issue observed in these interventions is their top-down approach, where end-users are not involved in the design and implementation process, leading to reduced sustainability, acceptability, and reliability of the interventions.

Human-centered design has demonstrated significant potential in generating highly acceptable and sustainable health solutions. This approach is particularly effective due to its capacity to introduce innovative solutions to entrenched challenges while incorporating the perspectives of multiple stakeholders, including end-users (21, 22). By capturing the unique needs and requirements of end-users, this method fosters a sense of ownership over the developed interventions, thereby enhancing their sustainability and leading to more positive outcomes (23). Considering knowledge barriers and feeding practices among caregivers of under five children that hinders achieving minimum food diversity, frequency and adequacy; this study therefore aims to co-design age specific meal plans based on locally available foods in Njombe and Iringa. By using a human-centered design approach, the research will involve caregivers in the co-creation process, ensuring that the resulting meal plans are culturally relevant, economically feasible, and practical for families from all socioeconomic backgrounds. This approach not only addresses the challenges caregivers face in providing age-appropriate nutrition but also promotes community ownership and long-term sustainability. Ultimately, the study seeks to improve nutritional outcomes for under-five children, reduce malnutrition rates, and contribute to broader public health goals, serving as a model for similar interventions in other regions.

## Methods

### Study design

This study will adopt a comprehensive five-step Human-Centered Design (HCD) approach, utilizing a qualitative descriptive design to co-design age-specific meal plans tailored to the needs of under-five children in the Njombe and Iringa regions of Tanzania. The HCD framework will facilitate active participation from all relevant stakeholders, ensuring that the solutions developed are culturally appropriate, sustainable, and grounded in local realities (24). The study will be conducted in five main steps: Exploration, Co-design, Validation and Refinement, Implementation, and Evaluation.

### Step 1: Exploration/ Community driven inquiry

During this phase, the research team will connect with study participants to empathize with them and understand the challenges they face in preparing age-specific meals for their children. Focus Group Discussions (FGD’s) and Key Informant Interviews (KII’s) will be used to gather information from caregivers of under-five children and other stakeholders (Regional nutritional officer, District medical officer, district reproductive and child health coordinator, district nutrition officer, and CHW’s). Their insights will guide the development of context-specific solutions.

### Step 2: Co-design phase/ Consultative meetings

This phase consists of three parts: synthesis, ideation, and co-design.

- **Synthesis:** Findings from Step 1 will be presented to participants to ensure all concerns raised during the community inquiry are captured accurately.
- **Ideation:** The focus will be on identifying locally grown foods or those available at lower prices, suitable for inclusion in meal plans.
- **Co-designing:** Locally available foods will be aligned with a pre-developed template indicating meal size, contents, and frequency appropriate for different age groups. This template was iteratively developed by the research team, which included specialists in nutrition.

### Step 3: Validation and Refinement

The emerging meal plan package will be validated through FGD’s with mothers and caregivers who did not participate in the previous two steps. Open-ended questions will explore whether the proposed interventions will help caregivers provide age-specific meals and gather their perspectives on the advantages, disadvantages, and any missing components. Based on these insights, the meal plan will be refined to ensure it is practical and well received by the community.

### Step 4: Implementation

The refined meal plan package will be implemented over one year in two regions: Njombe and Iringa. Community Health Workers (CHW’s) will be recruited and trained on the developed meal plan. They will identify families with under-five children and pregnant mothers, assess the nutritional status of the children, and provide continuous education to caregivers. CHW’s will play a key role in supporting the implementation and ensuring the plan is followed consistently.

### Step 5: Evaluation

In the evaluation phase the researchers will assess the effectiveness and sustainability of the meal plan intervention. The evaluation will occur multiple times throughout the one-year implementation period, using both qualitative and quantitative methods.

- **Quantitative evaluation:** The nutritional status of children will be assessed at baseline (before implementation), mid-point (6 months), and endpoint (12 months). Key indicators, including weight-for-age, height-for-age, and weight-for-height, will be measured to evaluate changes in nutritional outcomes.
- **Qualitative evaluation:** In-depth interviews and FGD’s will be conducted with caregivers, CHW’s, and key stakeholders to assess their experiences with the meal plan, challenges faced, and perceived benefits. This will also provide insight into the level of acceptance and adoption of the meal plan in the community.
- **Process evaluation:** The fidelity of implementation, including CHW’s’ adherence to training and the extent to which the meal plan is followed by caregivers, will be monitored. Regular supervision and feedback mechanisms will be established to ensure continuous support and improve the quality of implementation.

The findings from the evaluation will be used to refine the meal plan further and provide recommendations for scaling up similar interventions in other regions. Additionally, the evaluation will help identify any long-term changes in nutritional status and behavior within the targeted communities. Figure 1 provides a summary of the Human centered design steps starting from Exploration to Evaluation.

**Figure 1:**
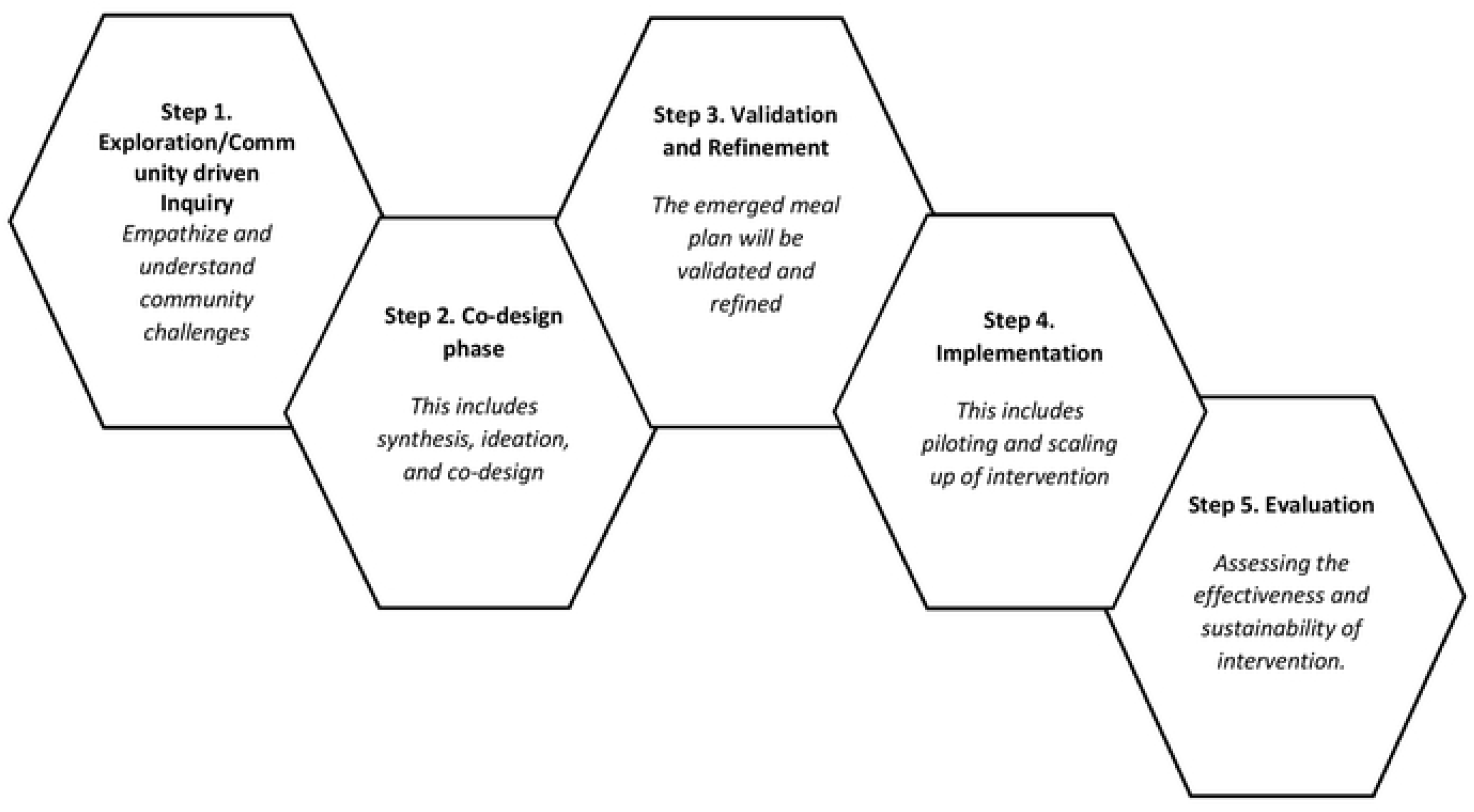
Five Human centered design steps.

### Study Area

The study will take place in Njombe and Iringa regions located in the Southern highland zones of Tanzania. Both regions are predominantly inhabited by the Bena, Hehe and Kinga ethnic groups. Other ethnic groups are Pangwa, Nyakyusa, Chagga but these are predominantly located in the urbanity of Iringa region for business purposes (25). Njombe Region consists of six administrative districts: Makambako Town, Ludewa, Njombe Town, Makete, Njombe, and Wanging’ombe; While Iringa on the other hand has 3 districts: Iringa district, Kilolo district and Mufindi district. These regions have been selected because; both Njombe and Iringa have been identified as regions with significant rates of malnutrition among under-five children (26). Tanzania demographic health survey 2022, indicated these regions with stunting prevalence more than 40% compare to other regions (6). Hence making them ideal locations to test and implement new approaches to child nutrition.

Moreover, Njombe and Iringa regions are characterized by a rich agricultural landscape, offering a wide variety of crops, fruits, and vegetables. Despite this agricultural wealth, there remains a significant gap in utilizing these locally available foods to improve child nutrition. By focusing on these regions, the study aims to tap into this agricultural diversity to develop meal plans that are not only nutritious but also locally sourced. This approach not only ensures the sustainability of the intervention but also creates a model that can be adapted and replicated in other regions with similar agricultural potential.

### Study Population

The target population for this study includes First: Primary caregivers of children under five years old, including parents, guardians, and extended family members, who are directly involved in the day-to-day feeding practices. Second: Community health workers (CHW), regional and district medical officers and nutritionists who provide health and nutrition services within the community and have expert knowledge of child nutrition. Third: Community leaders, elders, and traditional knowledge holders who possess an understanding of local food systems, cultural practices, and social dynamics.

### Sample Size, Sampling, and Data Collection

A total of 208 participants will be purposefully selected for this study. Data will be collected through focus group discussions (FGDs), key informant interviews (KIIs), and consultative meetings.

- **Step 1:** Twelve (12) KIIs and four (8) FGDs (each with a maximum of 8 participants) will be conducted in Njombe and Iringa, totaling 76 participants.
- **Step 2:** Two days consultative meetings will be held with 30 participants from each region, including caregivers of under-five children, community health workers (CHWs), nutritionists, community leaders, and regional and district medical officers.
- **Step 3:** The validation process will involve three (3) FGDs per region, each with up to 8 participants, totaling 48 participants. Only caregivers and stakeholders who have not participated in the previous steps will be included in this phase.

The final refinement process will take place indoors with the principal investigator (PI), Co-Principal investigators (Co-PI), and nutrition experts. The success of the above step will further allow implementation of the intervention and future evaluation.

### Data Management and Analysis

Data will be collected by four trained research assistants under the supervision of PI and Co- PI. Research assistants will handle simultaneous transcription and translation, with verification by the research team. All interview transcripts will be deidentified, and pseudonyms will be assigned to participants. The data will be uploaded into NVivo 12 software (QSR International) for organization and thematic coding.

A deductive thematic analysis will be conducted step by step. The research team will first review the research questions and collaboratively identify key themes, forming an analytical matrix of main themes and subthemes. Responses from participants (codes) will be systematically assigned to corresponding themes and subthemes within NVivo. If any codes do not align with the established themes, the research team will review them using a consensus-based approach, excluding those that do not contribute significant insights. The structured data will be transferred to Microsoft Word for in-depth interpretative analysis and the preparation of the final report.

### Ethics Approval

This research will obtain ethical approval from Agha Khan University (AKU) Ethics Review Committee and then the approval from the Presidency Office Reginal Administration Local Government (PORALG), along with local permissions from the PORALG, the permission from respective regional authorities will be secured. Verbal consent will be requested from local government, following the submission of official letters from PORALG and copies of the ethical clearances.

To ensure the ethical integrity of the study, informed written consent will be obtained from all participants prior to their involvement. The research will not involve any medical diagnosis or treatment, thus posing no direct or indirect risks to the participants related to health. All responses will remain confidential, with data analysis and reporting conducted in aggregate at the respected region. The collected data will be strictly used for the purpose of this study.

### Dissemination plan

Findings from this study will be disseminated through multiple channels to ensure they reach the appropriate stakeholders and the public for maximum impact on policy, education, and practice. The research findings will be presented to the Ministry of Health and Tanzania food and nutrition center. These two entities will be key in integrating the findings into national child nutrition policies and intervention programs. Relevant stakeholders, including non-governmental organizations (NGOs) and other entities involved in child welfare and nutrition, will be engaged through consultative meetings and workshops.

Moreover, the researchers plan to present findings at national and international health conferences, fostering engagement with healthcare professionals, academics, and policymakers to promote evidence-based practices and informed decision-making. Presenting at such forums will facilitate the sharing of knowledge with a wider audience, stimulate discussions around child nutrition, and help position the findings in global discourse. It will also provide an opportunity for peer feedback and further validation of the findings. To ensure the public is informed about the importance of child nutrition, key messages derived from the research findings will be disseminated through various media outlets, including newspapers, radio, television, and social media platforms. This will serve to educate parents, caregivers, and communities on best practices for child nutrition and raise awareness of the potential risks of malnutrition. The research will also be disseminated through academic journals and publications to contribute to the broader body of scientific knowledge on child nutrition. Research reports will be made available to educational institutions, promoting the integration of the findings into academic curricula and future research.

### Study Timeline

Table 1 below shows study timelines indicating the key phases and activities of the study from protocol development to full-scale implementation. It provides a structured timeline, indicating completed tasks and upcoming milestones to ensure smooth progress.

**Table 1:** Study timeline.

| Phase | Activity | Timeline | Status |
| --- | --- | --- | --- |
| Protocol Development and Review | Drafting study protocol | December 2024 – January 2025 | Done |
| Ethical Review Application | Submission of ethical review application | January 2025 | Done |
| Protocol Submission for publication | Review of Journal requirements and submission for publication | February 2025 | Done |
| Fundraising | Engaging partners and stakeholders to secure funding | February – October 2025 | Ongoing |
| Study Initiation | Securing approvals from government authorities | November 2025 | Not Done |
| Step 1: Community driven Inquiry | Initial participant recruitment, Focus group discussion, Key Informant's interviews and preliminary data analysis. | November 2025 | Not Done |
| Step 2: Co-design | Participants of step 1 above will be called for consultative meetings | December 2025 | Not Done |
| Step 3: Validation & Refinement | Second phase participants recruitment, Focus group discussions for validation and refinement meetings. | January – February 2026 | Not Done |
| Step 4: Implementation (Piloting Phase) | Conducting pilot study | March – December 2026 | Not Done |
| Step 5: Evaluation | Assessing pilot study outcomes | January – February 2027 | Not Done |
| Scale-up Phase | Expanding study implementation (subject to funding availability) | From March 2027 | Not Done |

## Discussion

This study embarks on an innovative approach to tackle childhood malnutrition by co-designing tailored, age-specific meal plans for under-five children in Iringa, Njombe, and Ruvuma. Age specific meal plans will serve as an intervention to address knowledge gaps among caregivers, ensuring children receive adequate nutrition in the appropriate quantities, frequencies, and with diverse food options. Unlike previous interventions aimed at combating malnutrition, which often adopted a top-down approach and overlooked cultural relevance, acceptability, and sustainability, this study will employ a human-centered design approach to ensure these factors are prioritized.

The co-designed meal plans will be presented in a weekly format, providing caregivers with a clear guide to feed their children a variety of locally available foods from Monday to Sunday. The plan will include a wide range of food options, including between-meal snacks, and specify recommended portion sizes based on the child’s age and the nutritional content of each food item. This intervention will not only address undernutrition but also contribute to preventing overweight and obesity among under-five children. To further support caregivers, user-friendly food measurement tools will be suggested, making the implementation of the meal plans practical and accessible.

The anticipated outcomes of this study include improved dietary diversity and feeding practices for under-five children. The research builds on existing evidence demonstrating that meal plans with a diverse range of foods can significantly improve a child’s nutrient intake, promoting healthy growth and cognitive development (27, 28). Given that poor infant and young child feeding (IYCF) practices are a major contributor to undernutrition in Tanzania, the proposed age-specific meal plans will provide a practical solution that addresses the unique needs of children at different developmental stages (29).

Furthermore, the study’s focus on locally available foods will ensure that the meal plans are not only nutritionally adequate but also economically viable for families across socioeconomic strata. This is critical, as economic constraints and food insecurity are among the primary factors contributing to childhood malnutrition in low- and middle-income countries like Tanzania (30). By focusing on affordable, locally sourced ingredients, the intervention has the potential to be scaled across similar settings within the region, ensuring broader impact.

However, the study also anticipates certain challenges in its implementation. While the focus on locally available foods enhances sustainability, seasonal variations could disrupt the consistency of meal planning for some families. To address this, the co-design process will need to incorporate flexibility and explore food preservation and storage techniques that ensure year-round nutritional support.

In conclusion, this research holds the potential to contribute meaningfully to reducing malnutrition among under-five children in Tanzania by creating practical, culturally relevant, and sustainable meal plans. The human-centered design approach, which emphasizes user participation, may serve as a model for future public health interventions aimed at improving child nutrition across Tanzania and similar regions. If successful, this study could provide critical insights for achieving Tanzania’s national nutrition goals and aligning with global initiatives such as the Sustainable Development Goals (SDG 3), which prioritize health and well-being for all ages.

## Data Availability

In request, the data can be requested through my

## Acknowledgements

We extend our deepest gratitude to the entire Health One on One Africa team for their unwavering support and collaboration throughout the proposal development process. Specifically, the invaluable contributions of Justice Komba, Romelly Edward, Vivian Sekei, and Yuda Sulle Pascal, whose dedication and expertise have been instrumental in shaping this work.

We also express our heartfelt appreciation to our colleagues for their insightful feedback and continuous encouragement. Our sincere thanks go to our families and friends, whose unwavering support and understanding have been a source of motivation throughout this journey. Their belief in our vision has been truly invaluable.

